# Breadth of humoral immune responses to the C-terminus of the circumsporozoite protein is associated with protective efficacy induced by the RTS,S malaria vaccine

**DOI:** 10.1101/2020.11.15.20232033

**Authors:** Sidhartha Chaudhury, Randall S. MacGill, Angela M. Early, Jessica S. Bolton, C. Richter King, Emily Locke, Tony Pierson, Dyann F. Wirth, Daniel E. Neafsey, Elke S. Bergmann-Leitner

## Abstract

The circumsporozoite protein (CSP) is the main surface antigen of malaria sporozoites and a prime vaccine target. Responses induced by the CSP-based RTS,S vaccine towards the polymorphic C-terminal region of *P.falciparum*-CSP raise concerns that vaccines using single alleles may have lower efficacy against genotypic variants. We characterized the extent of C-terminal cross-reactivity of antibodies induced by RTS,S (based on the 3D7 allele) with variants representing seven circulating field isolates through a novel HTS-multiplex assay for screening closely related peptides. Reactivity to variants showed approximately 30-fold reduction in recognition relative to 3D7. The degree of reduced cross-reactivity,ranging from 21 to 69-fold, directly correlated with the number of polymorphisms between variants and 3D7. Surprisingly, protection assessed by challenge with 3D7 parasites was strongly associated with higher C-terminal antibody breadth suggesting that C-terminal specific avidity or fine-specificity may play a role in RTS,S/AS01_B_-mediated protection and that breadth of C-terminal CSP-specific antibody responses may be a marker of protection.

## Introduction

Most malaria parasite antigens exhibit extensive population-level diversity, resulting from millennia of selection pressure to escape naturally acquired immunity [1]. The Circumsporozoite Protein (CSP) is the main surface antigen on the infecting sporozoite and has been targeted by a variety of pre-erythrocytic vaccines. The leading *Plasmodium falciparum* CSP-based vaccine is RTS,S/AS01_E_ which received a positive scientific opinion (under Article 58) from the European Medical Agency in 2015, and is currently subject to pilot implementation in three African countries with high malaria endemicity [2, 3].A study conducted in Malawi that focused on T cell epitopes characterized the naturally occurring variation in CSP and found evidence that naturally-acquired immunity provides selective pressure on the CSP C-terminus [4]. Further evidence of such immune pressure was provided by sequencing the CSP C-terminus polymorphisms of *P. falciparum* infecting unprotected RTS,S/AS01_E_-vaccinated children (ages 5-17 months). The genetic diversity of parasites in vaccinated subjects demonstrated that vaccination leads to improved protection against 3D7-matched parasites [5]. This implies vaccine efficacy could be partially dependent on matching the C-term allele(s) to those prevalent among parasites in the geographic region of vaccine deployment.

Polymorphism in the CSP C-terminus presumably reflects a balance between immunity driven selection and constraints to preserve protein function. The C-terminus of CSP is crucial for invading hepatocytes and, therefore, for successful infection of the liver and establishment of the disease [6]. Field data from a Phase III trial in African children and infants has demonstrated the importance of C-terminal responses in RTS,S/AS01_E_-mediated protection [7]. This study also identified polymorphisms within the C-terminus of the CSP that are associated with vaccine efficacy. The variant sequences associated with protection identified in clinical field samples [5] represent the foundation for the present study.

Previous studies have not found a consistent association between immune responses to the CSPC-terminus and vaccine efficacy. Antibodies targeting the C-terminus have been associated with phagocytic activity, but the phagocytic opsonization index was found to be negatively correlated with protection in a phase 2 RTS,S trial [8]. A recent systems serology study extended protective correlates to NK cell activity and other Fc-mediated activities [9]. A recent study of B-cell responses in malaria-naïve adults who were vaccinated with live sporozoites identified only 2 out of 215 antibodies that were specific to the C-terminus from subjects who were protected from subsequent challenge infections, and these antibodies did not exhibit neutralizing characteristics *in vitro* or confer protection in mice challenged with PfCSP transgenic *Plasmodium berghei* parasites [10]. However, analysis of IgG in plasma/serum from subjects in the RTS,S/AS01_E_ phase 3 trial found protection to be significantly correlated with IgG avidity to the CSP C-terminus, suggesting this region plays an important but complex role in vaccine-induced protection [7, 11].

We recently adapted an electro-chemiluminescence based (ECLIA) multiplex platform to test serological responses to closely related antigens without cross reactivity due to antigenic similarity and competition. Moreover, the assay platform has a very high sensitivity with exceptionally low inter- and intra-assay variability (manuscript submitted). It enables monitoring the reactivity of either vaccine-induced anti-CSP antibodies or naturally acquired antibodies induced to the currently known C-terminal variants of the CSP.

The objective of the current study was to determine whether the breadth of the humoral immune response to CSP C-terminus variants can predict vaccine efficacy and protection. Pre-immune *vs*. immune sera from an RTS,S Phase IIa clinical trial assessing protective efficacy in U.S. adults [12] were tested for reactivity against peptides representing CSP-variants identified in the above-mentioned field study [5]. In this study, volunteers received RTS,S formulated with AS01_B_ or AS02_A_, and no difference in serological responses was observed between the different formulations, consistent with pre-clinical studies [13]. The results indicate that protected individuals react against a wider range of peptides, i.e. breadth of response to variant peptides, compared to individuals who are not protected following controlled human malaria infection (CHMI). These results can be used to guide the refinement of the design of CSP-based malaria vaccines to develop a vaccine with increased breadth of reactivity to C-terminal variants, which may be a correlate of protection in RTS,S vaccinated subjects.

## Materials and Methods

### Peptides

Eight biotinylated PfCSP C-terminal peptides were produced by CS Bio (Menlo Park, CA) and provided by PATH (CSP aa 283-375). Sequences are listed in Table 1. This region of the CSP C-terminus spans most of the non-repeat region of the RTS,S vaccine construct (CSP aa 272-389) and harbors many highly polymorphic amino acid positions presumably selected by naturally acquired immune responses, and for which a subset were observed to be associated with allele-specific vaccine protection in a previous study [5].

### Antibodies

Pre-immune and day of challenge sera (two weeks post third immunization from a previously conducted clinical trial (NCT00075049) [12] in which study participants vaccinated with RTS,S adjuvanted in AS01_B_ or AS02_A_ (n =15 protected subjects, n =11 non-protected subjects) were tested for reactivity against C-terminal peptides. Preliminary experiments did not show differences between the two vaccine cohorts in their reactivity to the variant peptides. The serum sample use was reviewed by the WRAIR Human Subjects’ Protection Branch which determined that the research does not involve human subjects (NHSR protocol WRAIR#2142) as the samples used were de-identified and no link between samples and subjects exists. A human CSP-immune serum pool (CSP-AV) and commercial human AB pooled serum were used as positive and negative assay controls respectively.

### Mesoscale Diagnostics

The experiments were conducted using the U-PLEX assay platform (Mesoscale Discovery (MSD) Inc, Gaithersburg, MD) as previously reported [14]. In brief, individual biotinylated peptides were incubated with the proprietary U-PLEX MSD linkers designed to bind to respective spots on 10-spot MSD multi-array 96 plates. U-PLEX linker-coupled peptides were combined into a cocktail containing all eight peptides (*i.e*., H12, H13, H18, H234, H3, H50, Pf16-MVI, Pf16-H1; 300 nM of each pre-linked peptide). Plates were coated overnight at 4°C and washed three times with 1x Wash Buffer (MSD). Serum samples were diluted 1:5,000 with assay diluent (diluent 2, MSD) and incubated for 1 hour at RT on shaker. Plates were washed and incubated with goat-anti-human IgG (H+L) secondary antibody conjugated to Sulfotag (MSD, 1 µg/ml final concentration) in diluent 3 for 1 hour at RT on shaker. Plates were washed and 2x Read Buffer T (MSD) was added prior to reading the plates in the Meso Quickplex SQ120 per the manufacturer’s instructions. Data were reported as mean luminescence signal (MLS).

### B cell epitope predictions

Sequences of the variant and 3D7 C-terminus were used as the input for the sequence-based linear B cell epitope predictions using BepiPred [15]. The crystal structure deposited in the Protein Data Bank (PD ID code 3VDJ) [16] was used for predicting conformational epitopes in the 3D7 sequence with the Discotope method [17]. The tools were accessed through the Immune Epitope Database (www.iedb.org)[18].

### Statistics

Univariate analysis between protected and unprotected subjects was performed to determine the correlation between MSD intensity (reactivity) to the various peptides and protection. Prior to any t-test, we carried out a Shapiro-Wilks test to determine if the to-be-compared data points were normally distributed. If both were normally distributed (p < 0.05 by the Shapiro-Wilks test), we applied a Student’s t-test; if either distribution was not normally distributed, we applied the Wilcoxon signed-rank test.

To express the breadth of the immune response, first the relative response of individual sera to the different variant peptides was calculated (Px = MLS to peptide y/MLS to 3D7 allele) and then the median response across all tested variant peptides was calculated (breadth = median (P_3D7_, P_H1_, P_H33_, P_H12_, P_H50_, P_H13_, P_H18_, P_H234_). Binomial logistics regression was performed to estimate the probability of protectionbased on antibody breadth. Correlations and correlation matrices were calculated and plotted using R software. R script and data are available upon request.

### Data availability

All analysis scripts used in this study were written in R and are available freely for download at https://github.com/BHSAI/immstat. Experimental data and protocols are made available upon request to corresponding author.

## Results

### Design of C-terminal CSP variant peptides

Seven C-terminal peptides were selected from a database of naturally occurring CSP sequences observed in the RTS,S/AS01_E_ phase III ancillary genotyping study [5]. Each peptide contains varying degrees of similarity to the 3D7 vaccine construct at polymorphic amino acid positions, including those observed in the Phase III ancillary genotyping study to contribute to differential vaccine efficacy [5]. Previous studies of CSP have highlighted three regions hypothesized to serve as T-cell epitopes [19] and/or B-cell epitopes [20, 21]: DV10 (positions 293-302), Th2R (positions 311-327) and Th3R (positions 352-363). Figure 1 aligns the seven peptide constructs along the 3D7 sequence (peptide MVI-Pf16) and indicates highly polymorphic positions within the DV10, Th2R, and Th3R epitope regions. The magnitude of polymorphism observed in the phase III ancillary genotyping study [5] is represented by potentially balanced polymorphisms, indicating positions where the major allele is present with a frequency < 80%, and whether or not a position was a ‘sieve site’ (associated with differential vaccine efficacy) [5]. Three of the four positions with the highest sequence entropy occur in Th2R, specifically 318E, 321N, and 322K, along with one position (352N) in Th3R (not shown). The H18 peptide has the smallest Hamming distance to 3D7, with five mismatching amino acid positions; H50 is the most distant from 3D7 with 10 mismatches.

**Figure 1:**
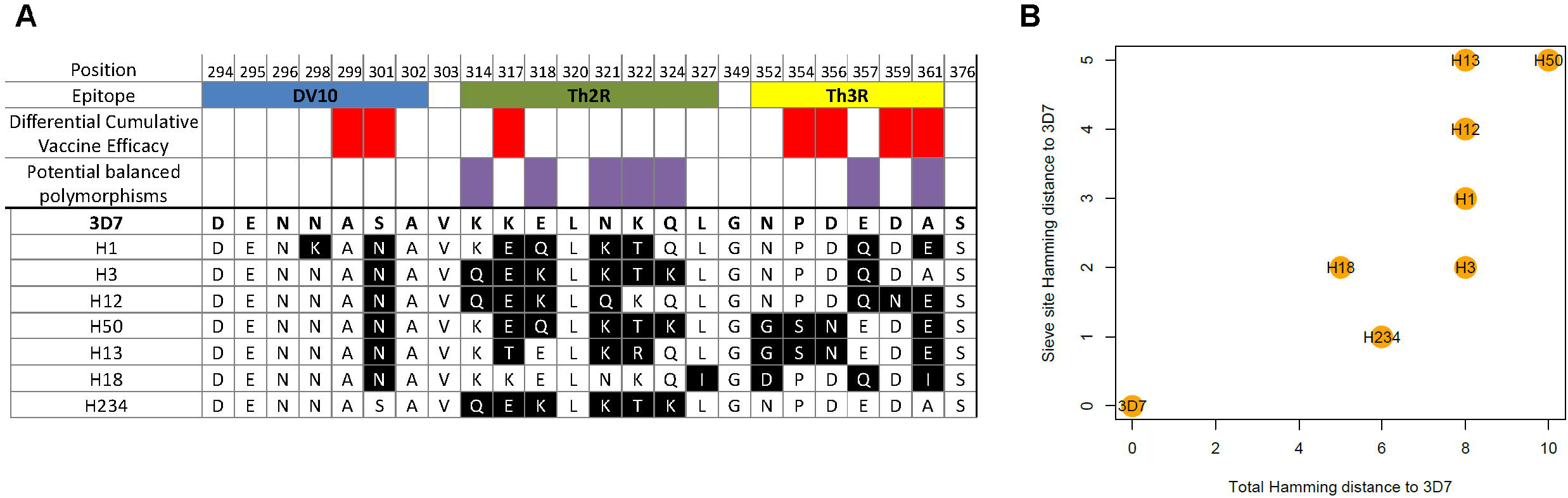
CSP-variant peptide sequence alignment and distance from 3D7. A) Multiple sequence alignment of the seven variant peptides, compared with 3D7, along the DV10, Th2R, and Th3R epitope regions. Mismatched residues are shown in black. Hamming distance to 3D7 is shown for each peptide variant along with information on differential cumulative vaccine efficacy and potential balanced polymorphisms. B) ‘Sieve site’ Hamming Distance and Total Hamming Distance of the CSP-variant peptides and 3D7.

### Reactivity of RTS,S-immune sera to variant C-terminal CSP peptides

Sera from RTS,S-vaccinated subjects were tested using an ECLIA-based multiplex platform (MSD) for reactivity against C-terminal CSP variants that represent prevalent parasite strains across Africa. As expected, we found reactivity to the 3D7 peptide higher than the variant peptides in all cases (Figure 2A, Supplementary Figure 1). We observed on average, reactivity to the CSP variants showed a 30-fold reduction in signal intensity relative to 3D7, ranging from as low as 21-fold reduction to as high as 69-fold reduction across the seven variants. Additionally, there was a relationship between the degree of reduction in signal relative to 3D7 and the Hamming distance between that peptide and 3D7. H234 and H18, for example, were matching or highly similar to 3D7 at one epitope and showed a 21-fold and 25-fold reduction in signal, respectively, while H50, which was the most divergent sequence from 3D7 had a 69-fold reduction. Interestingly, the two peptides with the weakest cross-reactivity, H13 and H50, also display the greatest divergence from the five critical sieve site residues identified by Neafsey et al [5].Both H13 and H50 contain K317E, P354S, D356N, A361E mutations, possibly impacting linear and/or conformational epitopes.

**Figure 2:**
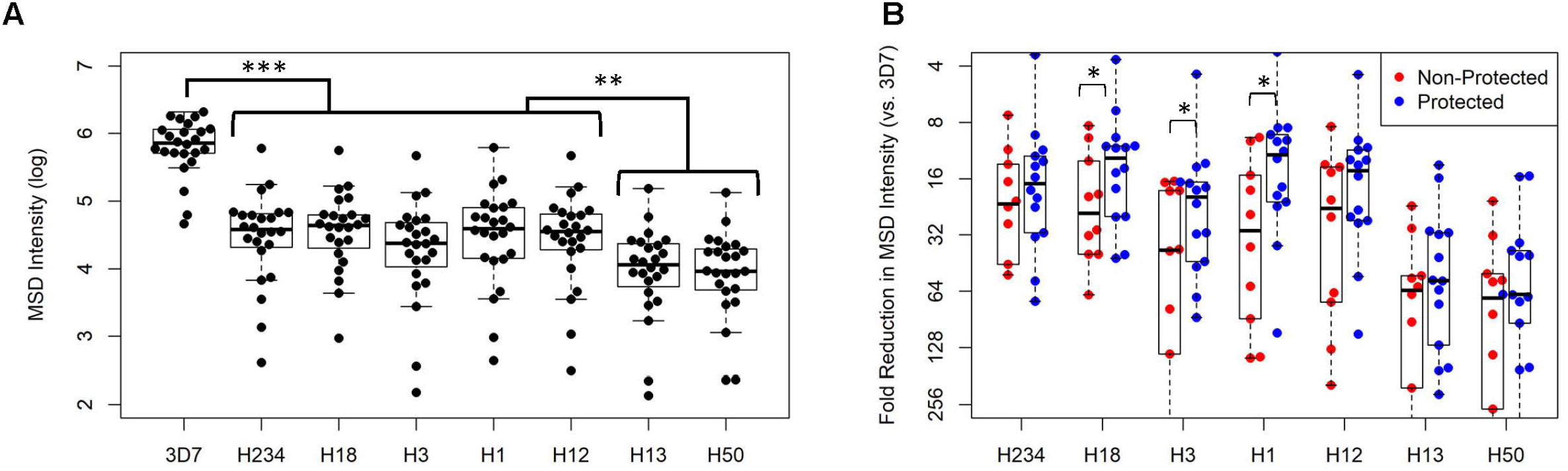
Serum reactivity of RTS,S-immune sera with 3D7 and seven variant CSP peptides. Dot plot summarizes reactivity of 26 vaccinees to the eight C-terminal peptides (A). Data expressed as net luminescence signal (signal of pre-immune sera subtracted; mean luminescence signal of CSP-negative sera 357 ± 120). Data were also stratified by protection status (blue for protected, red for non-protected) and shown as a fold change relative to 3D7 (B). Peptides are ordered in increasing sieve Hamming Distance, left to right, from 3D7. Peptides are ordered in increasing sieve-site Hamming Distance, left to right, from 3D7. Statistical significance shown with asterisks (* p < 0.05; ** p < 0.01; *** p < 0.001).

Stratifying the results based on protective status revealed (Supplementary Figure 2 and Figure 2B): 1) magnitude of the response to the 3D7 peptide did not discern between protected and non-protected individuals; (2) magnitude of the response to several peptides (H18, H12, H3, and H1) was significantly higher in protected individuals than non-protected individuals (p < 0.05 for H18, H12, and H3; p < 0.01 for H1). We also observed that this difference in protection status was mainly found in peptides with more modest differences from 3D7 – for example responses to H50 (Hamming distance of 10 from 3D7) showed no significant difference with respect to protection status, while responses to H18 (Hamming distance of 4) did. Overall among the peptides for which reactivity showed significant differences between protected and non-protected individuals, protected individuals showed approximately a 2-fold higher signal than non-protected individuals (Figure 2B).

### Breadth of CSP antibody response and protection

We sought to quantify the breadth of the antibody response in terms of its reactivity across *all* CSP variants in this study and assess its relationship to protection. We defined antibody breadth of a given sample as the median response across all the seven CSP variant peptides relative to its response to the 3D7 peptide. We compared this measure of antibody breadth by protection status (Figure 3A) and found that protected individuals had significantly higher breadth in their antibody responses than non-protected individuals (p < 0.01). We compared the antibody breadth relative to the antibody response to 3D7 to determine if the magnitude of the 3D7 response played a role in determining antibody breadth, but found no correlation between the two measures (Figure 3B). To further explore the association between breadth of antibody response to C-terminal peptides and protection, subjects were binned by antibody response breadth into three bins of equal width (Figure 3C). A binomial logistic regression to estimate the probability of protection from antibody breadth revealed that breadth is associated with the outcome (p=0.062), suggesting that the greater the breath of a subject’s antibody response, the more likely a subject is to be protected. The model suggests that a subject with low antibody response breadth (100-fold reduction in median signal between 3D7 and the CSP variants) has a 28% chance of protection while a subject with high antibody response breadth (10-fold reduction in median signal between 3D7 and the CSP variants) has a 84% chance of protection.

**Figure 3:**
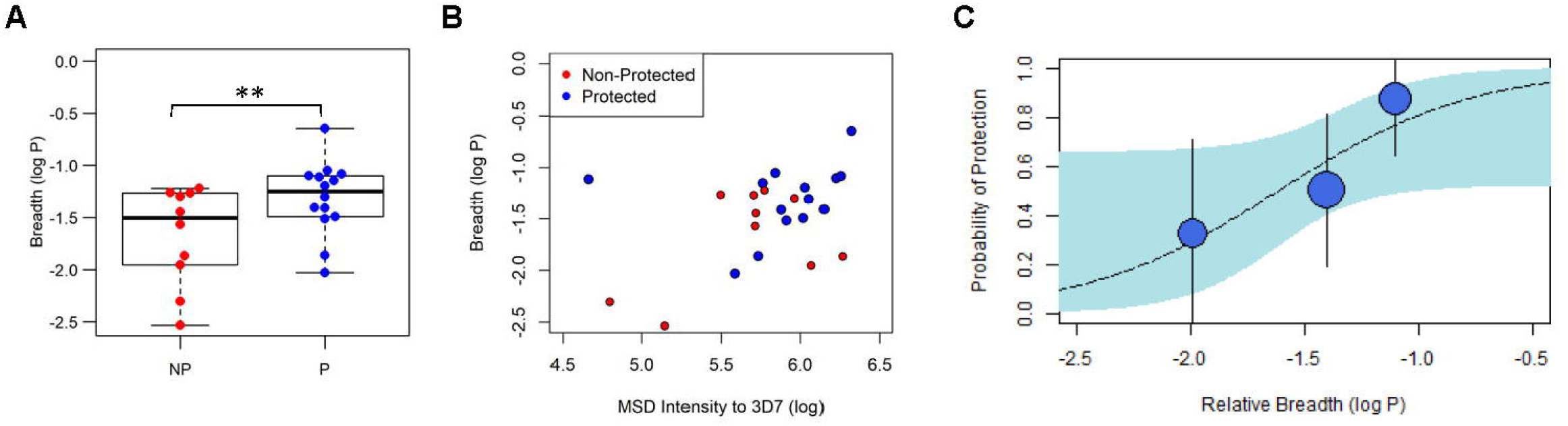
Breadth of the response to C-terminal CSP peptides is associated with protection. A) The breadth of the antibody response was expressed as median response across all tested variant peptides relative to 3D7. Data stratified into protected (blue) vs non-protected (red dots) (** p<0.01, Student’s t-test). B) Scatterplot showing breadth of antibody response compared to MSD intensity to 3D7; no significant correlation was seen in protected or non-protected subjects. C) Logistic regression was carried out with subjects binned into three bins based on relative breadth (circle size corresponds to sample size in bin; whiskers indicate 95% confidence interval). Estimated probability of protection based on antibody breadth shown with 95% confidence interval in blue.

Given that the overall breadth shows an association with protection, we sought to determine whether reactivity of the serum to pairs of peptides could provide additional insight. Using a correlation analysis, we identified peptides that show high correlation (i.e. subjects that show a high response to one peptide also show a high response to another or vice versa), suggesting that the antibodies in that sample are binding to epitopes in a cross-reactive manner between the two peptides. Conversely, peptides that show low correlation (i.e. a subject shows a high response to one peptide but a low response to another) suggests that antibodies in that sample are binding to epitopes in an allele-specific manner. Cross-reactivity in the antibody response can arise from two factors: 1) binding of antibodies to conserved epitopes and 2) binding of antibodies to conserved positions of otherwise largely polymorphic epitopes.

For protected subjects, we found that all peptides showed high correlation with each other (p< 0.05, Pearson correlation), suggesting that for these subjects, the antibody response appears to be cross-reactive across all eight peptides (Figure 4A). By contrast, for non-protected subjects we observed weak correlation between responses to 3D7 and the other CSP variants and a moderate correlation between all non-3D7 CSP variants, suggesting that their responses are largely allele-specific for 3D7. In both protected and non-protected subjects, the correlation between responses to all non-3D7 variants may reflect binding to conserved epitope positions. For protected vaccinees, this correlation extends to 3D7 as well, indicating in these subjects, RTS,S elicited responses that predominantly target conserved epitope positions. By contrast, for non-protected vaccinees, the lack of correlation between 3D7 and the other variants suggests that for these subjects, RTS,S-elicited responses target polymorphic epitope positions that are unique to 3D7 and exhibit high (albeit non-protective) 3D7 allele specificity.

**Figure 4.**
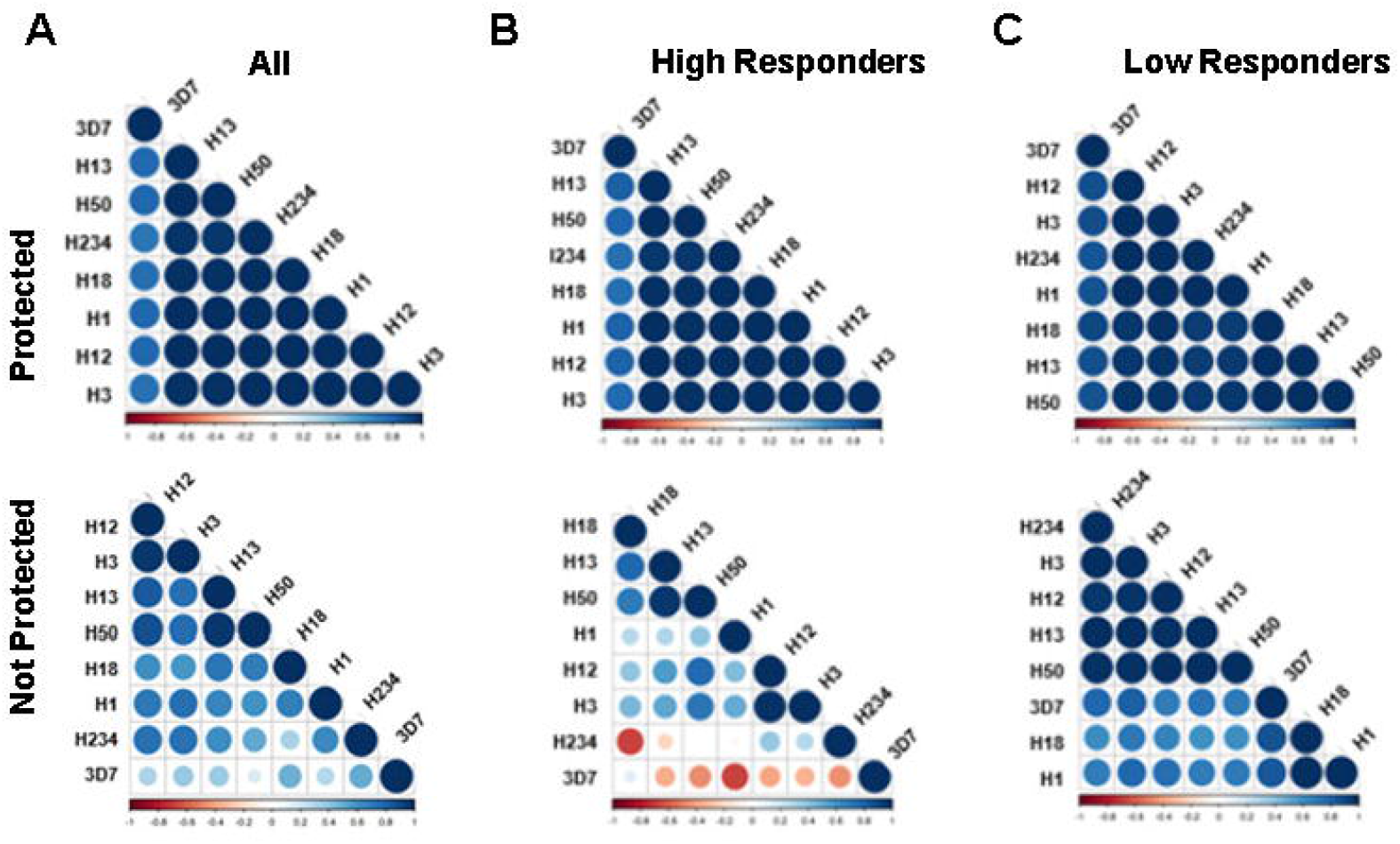
Antibody profile of protected vs. non-protected RTS,S immunized vaccinees to C-terminal peptides. Correlation matrices indicate the relationship between the magnitudes of the antibody responses to the various C-terminal peptides. The color and size of the dots (scale below graphs) indicate the degree of correlation between the different CSP peptides (small to large indicating low to high correlation). Correlation matrices stratified by protected status for all subjects (A), and separately subjects with high antibody titers (‘high responders’) (B) and subjects with low antibody titers (‘low responders’) (C). CSP-variant peptides are arranged based on clustering analysis to group peptides with similar patterns of responses together.

Supplementary Figure 3 shows representative cases for peptides H3 and H13 to illustrate this effect. In a comparison of responses to 3D7 and H3 across protected and non-protected subjects (Supplementary Figure 3A), protected subjects show a strong correlation (R^2^ = 0.57, p < 0.01) while non-protected subjects do not (R^2^ = 0.14). Likewise, in a comparison of responses between 3D7 and H13 (Supplementary Figure 3B), protected subjects also show a strong correlation (R^2^ = 0.60, p < 0.01), while non-protected subjects do not (R^2^ = 0.13). By contrast, in a comparison between responses between two non-3D7 alleles, H3 and H13 (Supplementary Figure 3C), protected subjects show a very high correlation (R^2^ = 0.96, p < 10^−3^) and non-protected subjects show a correlation as well (R^2^ = 0.65, p < 0.05). One interpretation for these results is 1) the correlation between the non-3D7 alleles and 3D7 in protected subjects suggests a significant portion of the RTS,S-induced response in these subjects is cross-reactive and 2) the correlation between responses to non-3D7 alleles suggests that cross-reactive portion of the RTS,S-induced response is broadly cross-reactive across non-3D7 alleles.

Next, the correlation analysis was stratified based on the overall ELISA titer to the 3D7 C-terminus (*i.e*., high titer *vs*. low titer in both, protected and non-protected groups) (Figure 4B and 4C). The stratification based on antibody titer and protection status revealed that antibody titers are not responsible for the distinct antibody reactivities against C-terminal variant peptides. Furthermore, we see distinct differences between high-titer and low-titer non-protected subjects. For low-titers, there is a high correlation across all the peptides, suggesting that responses in these subjects are targeting conserved epitope positions across these peptides. However, non-protected subjects with high C-term titers show a strikingly different pattern with a *negative* correlation between 3D7 and the non-3D7 alleles. This indicates for these subjects a stronger 3D7 response corresponded to a *weaker* non-3D7 response, suggesting non-protected subjects with high C-term titers are characterized by strong antibody responses towards 3D7-specific epitopes.

### Epitope prediction and structural modeling

Given the potential role of fine specificity in determining antibody breadth and its association with protection, we sought to characterize the antibody epitopes on the CSP C-terminal region. We carried out sequence- and structure-based epitope prediction (Figure 5A). The C-terminal region of CSP consists of a disordered linker region that connects the C-terminal region with the NANP repeat region and an ordered thrombospondin-like repeat domain unique to CSP termed the α-thrombospondin repeat (TSR) domain. We carried out linear epitope prediction on the linker region using the BepiPred algorithm [15] and found a major epitope region from positions 284-311. This region includes the DV10 epitope and has been previously reported to be an epitope recognized by sera from individuals immunized with irradiated sporozoites as well as by sera from children living in malaria endemic areas [20].

**Figure 5.**
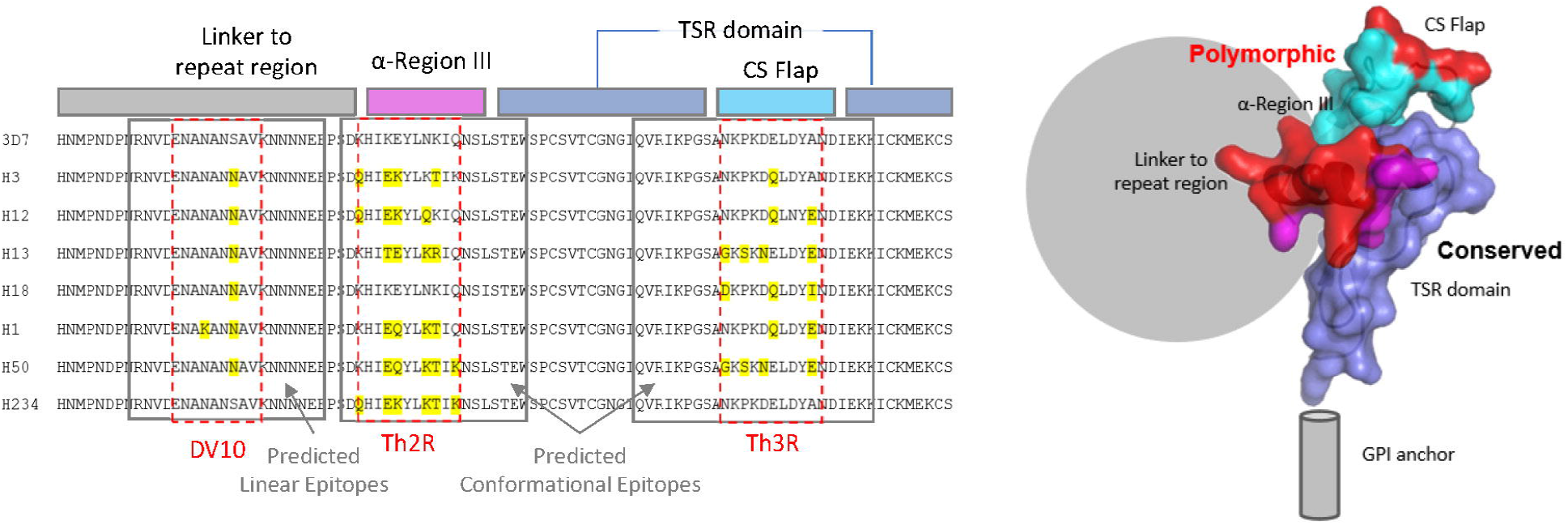
Sequence alignment, domain map, and structure of the C-terminal region of CSP. A) Sequences of CSP variants are aligned with 3D7 with polymorphisms highlighted in yellow and the DV10, Th2R and Th3R regions bound in red. The domain map of CSP C-term is shown with the linker region (gray), α-Region III (magenta), CS-flap (cyan), and α-TSR domain (dark blue). B) The structure of the α-TSR domain of CSP (PDB: 3VDJ) is shown in colors that correspond to the domain map, with polymorphic sites highlighted in red. Predicted epitope regions are shown in a ‘surface’ depiction on the structure.

We used the crystal structure of the α-TSR domain [16] of CSP for structure-based epitope prediction using the DiscoTope algorithm [17]. The algorithm identified two epitope regions – residues 311-331 and residues 342-367. These conformational epitopes correspond to the polymorphic Th2R and Th3R, respectively. The epitope containing the Th2R region (AA311-331) has been previously described as a B cell epitope in humans, rodents, and nonhuman primates providing some validation of these predictions [22, 23]. We mapped the epitope regions onto the structure of the CSP C-terminal region (Figure 5B). The Th2R and Th3R polymorphic regions map onto the two unique aspects of the CSP α−TSR domain – the α-helix in region III and the CSP ‘flap’, respectively. The α-TSR domain itself is highly conserved. Overall, based on the structural modeling there appear to be three overlapping epitope regions – the polymorphic α-helix region (containing Th2R), the polymorphic CSP-flap (containing Th3R), and the conserved ‘back-face’ of the α-TSR domain.

Given the potential role of conserved epitopes or epitope positions in determining antibody breadth, our structural modeling suggests that the major conserved epitope region lies along the back-face of the α-TSR domain, while polymorphic epitopes are primarily found on the ‘front-face’, along the α-helix of Region III and the CSP-flap that make up the Th2R and Th3R polymorphic regions, respectively. Our results suggest that, responses in non-protected subjects with high C-term titers may be mostly focused on these polymorphic epitopes.

## Discussion

While antibody responses to the CSP central repeats are known critical mediators of RTS,S/AS01_E_ vaccine efficacy, this is the first report revealing the relationship between breadth of antibody responses to different CSP C-term variants and protection from *P.falciparum* infection following RTS,S immunization. There are several potential strategies to improve the RTS,S/AS01_E_ vaccine efficacy and extend the durability of protection. First, efficacy could be boosted by broadening the immune responses to the C-terminus of CSP, which is known to be crucial for the function of the parasite and may contribute to increased vaccine efficacy. CSP can assume two different conformations: the non-adhesive conformation during migration of the parasite to the target tissue (i.e., salivary gland in the mosquito and liver in the mammalian host) and the adhesive form when preparing for invasion [6, 24]. When assuming the non-adhesive conformation, the N-terminus of the CSP folds over the C-terminus thus shielding it from potential immune attacks (such as antibodies or complement [25]). Most epitopes of the C-terminus are only accessible for antibody binding when the CSP assumes its adhesive conformation [24].Analyzing the differences between these two forms of CSP reveals the importance of the C-terminus for invasion, as the N-terminus of the CSP will protect this region during the sporozoite’s journey to its target tissue. Reports investigating the relationship between the epitope specificity of C-terminal monoclonal antibodies (mAbs) and their functional activity have shown that mAbs binding to the α-TSR have limited functional activity, likely because most epitopes are masked by the N-terminus (non-adhesive conformation) unless the parasite prepares for host cell invasion [10, 26]. In contrast, all tested mAbs binding to repeat and C-terminus have been shown to be invasion-inhibitory, while mAbs binding only to the α-TSR have limited functionality [26]. Selective immune pressure may be responsible for the increased frequencies of polymorphisms in the C-terminus, which is a balancing act between immune escape and maintaining functionality. The present study provides an insight into the breadth of C-terminal antibody specificities induced by RTS,S, given the tested samples were from malaria-naïve RTS,S vaccinees. The results also inform the potency of the vaccine in priming cross-reactive responses to other CSP C-terminal variants. Overall, we found that signal intensities corresponding to antibody titers were∼20-to 70-fold lower in the vaccine-mismatch alleles compared to the vaccine-matched 3D7 allele. While antibody responses clearly play a critical role in RTS,S-mediated protection, it is not known what consequences this reduction in reactivity in the C-terminal CSP response has for protection against diverse *P.falciparum* strains found in malaria endemic regions.

As has been the case in previous RTS,S studies [7, 8], there was not a significant difference in antibody titers to the C-terminal region of CSP between protected and non-protected subjects. However, interestingly, we did find that serum antibodies from protected subjects had higher reactivity to several CSP variant peptides than non-protected subjects. Furthermore, using a measure of antibody breadth based on the median response across all CSP C-terminal variants, protected subjects showed greater breadth than non-protected subjects. Logistic regression revealed a possible quantitative relationship between breadth and protection – subjects with only a ∼10-fold decrease in reactivity to the CSP variants compared to 3D7 had a >80% chance of protection, while subjects that showed greater than 100-fold decrease in reactivity to the CSP variants had only a 28% chance of protection. This link between antibody breadth and protection is puzzling given that the subjects were exposed to a homologous challenge, for which antibody breadth alone would play no obvious role in protection.

One possible explanation is that apparent breadth of the antibody response may be a proxy for fine-specificity – or relative response to different CSP C-terminal epitopes. An antibody response that predominantly targets conserved epitopes would be expected to show strong antibody breadth across all CSP variants, while an antibody response that primarily targets polymorphic epitopes would be expected to show allele-specific behavior, and correspondingly, weak breadth. Using correlation analysis between the antibody reactivities to the CSP variant peptides, we showed that protected subjects had a strong correlation across all the CSP variant peptides, suggesting their antibody responses were focused primarily on conserved epitope positions. By contrast, non-protected subjects, and particularly non-protected subjects with high CSP titers, show low or even negative correlation between their 3D7 response and their response to the CSP variants, suggesting that in these subjects, RTS,S is eliciting responses predominantly towards polymorphic positions of the 3D7 CSP.

Using structural modeling of the α-TSR domain of the C-terminal region of CSP, we show that the polymorphic epitope regions, Th2R and Th3R, are primarily found in the α-helix in Region III and the ‘CSP-flap’ [16]. Interestingly, these structural features of the α-TSR region of CSP are unique to the *P. falciparum* protein and not found in any other α-TSR or α-TSR-like domain structure [16], suggesting possible functional significance or selective pressure. Epitope prediction in conjunction with structure modeling shows that polymorphic epitopes of CSP C-term are primarily found on the same ‘front-face’ of the protein, near the linker region that connects the α-TSR domain to the NANP repeat region, while conserved epitopes along the highly conserved α-TSR domain are primarily found on the opposite face, or ‘back-face’. Given this arrangement of conserved and polymorphic epitope positions along the CSP structure, it is possible that antibody responses with differing degrees of cross-reactivity have different fine-specificities with respect to which epitope regions they bind to.

Although antibody responses to CSP have long been recognized as important for protection against infection, the work reported here represents the first systematic study to dissect the association between protection and fine specificity of antibody responses (*i.e*., potential epitopes in the C-terminus) to this region as has been reported for CSP-repeat specific antibodies [27, 28]. The role of C-terminal antibodies has been debated in the literature, with some reports demonstrating the importance of C-terminal antibodies in protection [7, 29] while others have shown a lack of functionality [10]. Our findings suggest that the conserved TSR region of the C-terminus may contain key protective epitopes that may be of great interest to vaccine design, especially in light of the possibility that antigenic mismatch due to polymorphisms in the CSP C-terminal region may limit RTS,S vaccine efficacy [5, 30]. Finally, our study suggests that the overall breadth of the antibody response across CSP C-terminal variants may be a marker for protection in CSP-based vaccines.

## Data Availability

All analysis scripts used in this study were written in R and are available freely for download at https://
github.com/BHSAI/immstat. Experimental data and protocols are made available upon request to corresponding author.

## Abbreviations

AS: Adjuvant system (GSK nomenclature)
CHMI: Controlled human malaria infection
CSP: Circumsporozoite protein
ECLIA: Electro-chemiluminescence
MLS: Mean luminescence signal
MSD: Mesoscale Diagnostics
NK: Natural killer cells
RTS,S: Scientific name of malaria vaccine indicating the vaccine components
Th_R: Helper T region
TSR: Thrombospondin repeat

## Acknowledgements

The authors would like to thank CPT Jennifer Kooken, Ms. Tanisha Robinson, and Ms. Elizabeth Duncan for facilitating work at various stages, and reviewers at GSK including Opokua Ofori-Anyinam, Lode Schuerman, Giuseppe Chiapparo, Erik Jongert, and Sophie Racine for their valuable comments.The clinical trial NCT00075049 was sponsored by the US Army Medical Research and Development Command in collaboration with Walter Reed Army Institute of Research (WRAIR) and GlaxoSmithKline Biologicals SA (GSK). GSK was provided the opportunity to review a preliminary version of this manuscript for factual accuracy, but the authors are solely responsible for final content and interpretation.

## Authors’ contributions

JB performed the experiments and SC performed the statistical analysis. EL, CRK, RM, AE, DW, DN provided critical reagents and information for the study. EB-L designed the experiments; EB-L and SC compiled the manuscript. All authors contributed by scientific discussions and input. All authors reviewed and edited the manuscript.

## Declarations

EB-L, TP, and SC are government employees. Title 17 U.S.C. § 105 provides that “Copyright protection under this title is not available for any work of the United States Government, but the United States Government”. Title 17 U.S.C. § 101 defines US Government work as “work prepared by a military service member or employee of the US Government as part of that person’s official duties”.

## Disclaimer

Material has been reviewed by the Walter Reed Army Institute of Research. There is no objection to its presentation and/or publication. The opinions or assertions contained herein are the private views of the authors, and are not to be construed as official, or as reflecting the views of the Department of the Army or the Department of Defense. The investigators have adhered to the policies for protection of human subjects as prescribed in AR 70-25. This paper has been approved for public release with unlimited distribution.

